# Development and validation of an algorithm to identify severe sepsis onset from electronic medical records

**DOI:** 10.1101/2025.10.02.25336096

**Authors:** Ramin Homayouni, Shane Morrell, Joel Karsten, Riya Chahbra, Paul D. Bozyk

## Abstract

**Objective:** To develop and evaluate an automated algorithm to identify sepsis onset from the electronic medical records (EMR), referred to as time-zero (t_0_), to enable more accurate surveillance, quality improvement, and training related to SEP-1 aligned care.

**Materials and methods:** We developed an algorithm to construct a comprehensive timeline of systemic inflammatory response syndrome (SIRS) criteria and organ dysfunction (OD) using structured data, and documentation of infection (DOI) using both structured data and unstructured clinical notes. The algorithm scans each timeline to detect the co-occurrence of SEP-1 components within a 6-hour window to determine t_0_. Algorithm performance was assessed using 2,030 manually abstracted adult sepsis cases from a large academic health system in southeast Michigan.

**Results:** Using a subset of 516 abstracted cases, we show that including clinical notes achieves higher concordance of DOI with abstractors t_0_ (41.9%) compared to using antibiotic (27.9%) or culture order proxies (34.7%). Combining all three sources achieves the highest DOI concordance (44.4%). On average, the algorithm DOI time was significantly earlier than abstractors (mean: -0.30 h, 95% Cl: -0.51 to -0.09) across all 2,030 cases, resulting in a significantly earlier t_0_ (mean: -0.51 h, 95% CI: -0.65 to -0.36).

**Discussion:** Automated approaches to analyze EMR data offer a scalable framework for SEP-1 monitoring, research, and quality improvement.

**Conclusion:** Incorporating unstructured clinical notes improves detection of infection suspicion and enhances concordance with manual abstraction of sepsis onset.

## INTRODUCTION

Sepsis is characterized as a life-threatening organ dysfunction caused by an unregulated host response to infection. Recent data shows that in the United States approximately 1.7 million adult hospitalizations and at least 350,000 deaths annually are related to sepsis. In the hospital setting, more than one-third of the deaths involve sepsis.^1,2^ Early identification and timely care are therefore central to quality improvement efforts.

In 2015, the Centers for Medicare & Medicaid Services (CMS) introduced the Severe Sepsis and Septic Shock Early Management Bundle (SEP-1) as a publicly reported measure within the Hospital Inpatient Quality Reporting (IQR) Program.^3,4^ SEP-1 operationalizes time-bound elements such as, initial lactate measurement and re-measurement, blood cultures before antibiotics, broad-spectrum antibiotics, and fluid resuscitation into composite 3- and 6-hour bundles. While SEP-1 has focused hospital attention on sepsis care, evidence regarding its association with improved outcomes is varied. Large multi-hospital evaluations of SEP-1 implementation found increases in lactate testing and some changes in treatment patterns, but no associated improvement in short-term mortality.^5,6^ These findings, together with concerns about measure complexity and potential unintended consequences, have fueled ongoing debate about how best to drive better sepsis outcomes.

Concurrently, national guidance emphasizes the need for robust hospital sepsis programs capable of surveillance, feedback, and targeted training across care settings. The CDC’s Hospital Sepsis Program Core Elements and National Healthcare Safety Network survey data highlighted that many hospitals rely on electronic medical record (EMR)-generated alerts and predictive models to assist in rapid sepsis identification.^1,2^

Prior work has leveraged automation to define sepsis t_0_ for SEP-1 evaluation using structured data, such as physician orders for blood culture or antibiotics. Order-based proxies underlie widely used electronic surveillance definitions (eg, CDC Adult Sepsis Event), which define “presumed serious infection” by culture orders plus antibiotic administration near the time of organ dysfunction.^7,8^ For example, Baghdadi et al. (2020) used an automated algorithm to mark t_0_ when Sepsis-3 physiologic criteria were first met including clinician suspicion of infection based on orders for antibiotics, antifungals, or clinical cultures within a 48-hour window.^9^ However, prior studies have shown substantial variability in determining SEP-1 t_0_ because it relies on specific note phrases to document suspicion of infection by a physician.^7^ This emphasizes the need for including clinical notes that may better document the onset of sepsis.

The goal of this project was to develop and evaluate an automated algorithm to identify sepsis t_0_ retrospectively from electronic medical record (EMR) using both structured data and information extracted from clinical notes.

## METHODS

### Study Population

A total of 2,888 cases were initially abstracted for evaluation. Of these, 41 cases were excluded because sepsis t_0_ could not be determined by abstractors, and 740 were excluded because t_0_ could not be identified by the algorithm, leaving 2,030 cases for analysis. After removing outliers using the interquartile range (IQR × 3) method, subsets of cases with complete data were used to compare algorithm performance with manual abstraction for each SEP-1 criterion: t_0_ (n = 773), SIRS (n = 605), organ dysfunction (n = 606), and documentation of infection (DOI; n = 915).

### Definition of severe sepsis and septic shock

Sepsis onset (time-zero) was defined according to the CMS criteria for severe sepsis or septic shock.^3,4^ The criteria for severe sepsis is met when the following three conditions were met within a 6-hour window: (1) evidence of infection or suspected infection, (2) fulfillment of at least two systemic inflammatory response syndrome (SIRS) criteria [temperature >38.3°C or <36°C, heart rate >90 beats/min, respiratory rate >20 breaths/min, or white blood cell count >12,000/µL, <4,000/µL, or >10% bands], and (3) evidence of acute organ dysfunction. Organ dysfunction was defined by at least one of the following: lactate >2 mmol/L, hypotension (systolic blood pressure <90 mmHg, mean arterial pressure <65 mmHg, or systolic blood pressure decrease >40 mmHg from baseline), acute respiratory failure requiring invasive or non-invasive mechanical ventilation, creatinine >2 mg/dL, urine output <0.5 mL/kg/hr for ≥2 hours, bilirubin >2 mg/dL, platelet count <100,000/µL, or coagulation abnormalities (INR >1.5 or aPTT >60 seconds). Chronic conditions or medication-related abnormalities were not considered as qualifying evidence of organ dysfunction. The criteria for septic shock is met if it is explicitly stated in the encounter note or when severe sepsis is accompanied by either (1) serum lactate ≥4 mmol/L or (2) persistent hypotension within 1 hour following a 30 mL/kg crystalloid fluid bolus.

### Sepsis t_0_ algorithm

The workflow for the sepsis t_0_ algorithm is shown in Figure 2. Sepsis cases were identified using the following inclusion criteria:

- Inpatient or ED encounters
- Age >= 18
- Primary or secondary billing ICD-10 coded with any of the following: A02.1, A22.7, A26.7, A32.7, A40.0, A40.1, A40.3, A40.8, A40.9, A41.01, A41.02, A41.1, A41.2, A41.3, A41.4, A41.50, A41.51, A41.52, A41.53, A41.59, A41.81, A41.89, A41.9, A42.7, A54.86, R65.20, R65.21

As indicated in Figure 2, the data extraction is de-coupled from the algorithm, enabling the algorithm to process data from any source. In this implementation, the structured data and unstructured notes were extracted from the Epic Clarity database across the entire duration of the inclusion encounters. The output of the extraction process was a combination of CSV and plain text files which served as the input to the algorithm.

The algorithm consists of three main components. First, all clinical notes for all encounters were processed using a set of manually refined regular expressions (available upon request) to identify septic shock, severe sepsis, or physician documentation of suspected infection. Second, outputs from the structured data fields and regular expressions were integrated to generate an encounter-specific event timeline. Finally, as depicted graphically in Figure 3 and detailed in the pseudocode (Figure 4), the algorithm evaluated each encounter timeline to determine whether the three t_0_ criteria were satisfied within a rolling 6-hour window. An example timeline is presented in Supplementary Figure S1.

## RESULTS

### Proxies for documentation of infection

Detection of infection documentation in EMR is challenging because it requires analysis of unstructured clinical notes. Prior studies have relied on proxy markers of infection, such as antibiotic or culture orders.^9^ In this study, we compared the algorithm’s ability to identify t_0_ using a variety of sources with the t_0_ documented by the human abstractors (Figure 5). The algorithm exactly matched the abstractor-recorded t_0_ in 41.9% of cases. When culture orders or antibiotic administration was used separately as a proxy for suspected infection, concordance with abstractor t_0_ was 34.7% and 27.9%, respectively. Combining all three sources achieved a concordance of 44.4%, which was marginally better than using the clinical notes alone. Notably, all three sources separately or combined identified t_0_ earlier than abstractors, ranging from 26.9% to 38.4% of cases.

### Evaluation of algorithm performance

Next, we examined the performance of the algorithm for each component of the SEP-1 t_0_ criteria (Figure 6). On average, there was no significant difference (mean: -0.05 h, 95% CI: -0.21 to 0.11) in the time of organ dysfunction identified by the algorithm and human abstractors (Figure 5). In contrast, SIRS time was significantly later (mean: 0.13 h, 95% CI: 0.03 to 0.23) by the algorithm. Notably, the algorithm identified DOI significantly earlier (mean: -0.30 h, 95% CI: -0.51 to -0.09) than the abstractors, resulting in a significantly earlier t_0_ (mean -0.51 h, 95% CI; - 0.65 to -0.36).

## DISCUSSION

Our findings suggest that defining sepsis t_0_ with an automated, EMR-based algorithm that incorporates both structured data and clinical notes can identify sepsis more accurately than structured data alone. In this study, the algorithm identified t_0_ earlier or at the same time as abstractors when documentation of infection (DOI) was based on clinical notes compared with culture or antibiotic orders (72.3% vs 61.6% and 50.0%, respectively). While proxies such as culture or antibiotic orders capture most cases, augmenting them with information from clinical notes produces more realistic t_0_ determinations at scale.

Large multi-hospital evaluations have reported increased lactate testing and changes in treatment patterns after SEP-1 implementation but no improvement in short-term mortality.^5,6^ However, these studies operationalized t_0_ and suspicion of infection primarily from structured EMR data (e.g., blood culture or antibiotic orders). Repeating such analyses with t_0_ definitions that incorporate free-text documentation could clarify whether measure timing, rather than the bundle itself, contributes to the null findings.^9^ If earlier, documentation-informed t_0_ demonstrates stronger associations between timely care and outcomes, that would support refining both surveillance and quality measures toward more clinically grounded anchors. Earlier t_0_ is clinically meaningful, as delays in antibiotic therapy and completion of early bundle elements are associated with higher mortality.^10,11^

In our study, using clinical notes led to significantly earlier identification of sepsis t_0_ compared with abstractors. This finding reflects that suspicion of infection is often documented before orders for cultures or antibiotics are placed. Earlier detection also partly reflects the algorithm’s ability to overcome human error. For example, Supplementary Figure 1 illustrates a 4.5-hour discrepancy between algorithm- and abstractor-determined t_0_, which resulted from missed documentation in a note. Such errors are expected when manual abstraction requires review of large volumes of clinical text. From an operational standpoint, our work demonstrates a path toward more efficient core measure reporting. SEP-1 abstraction is labor-intensive and vulnerable to inter-abstractor variability, particularly around time-zero determination.^7^ Having an algorithm-derived timeline for each sepsis case could improve both the efficiency and accuracy of human abstraction.

**Figure 1.**
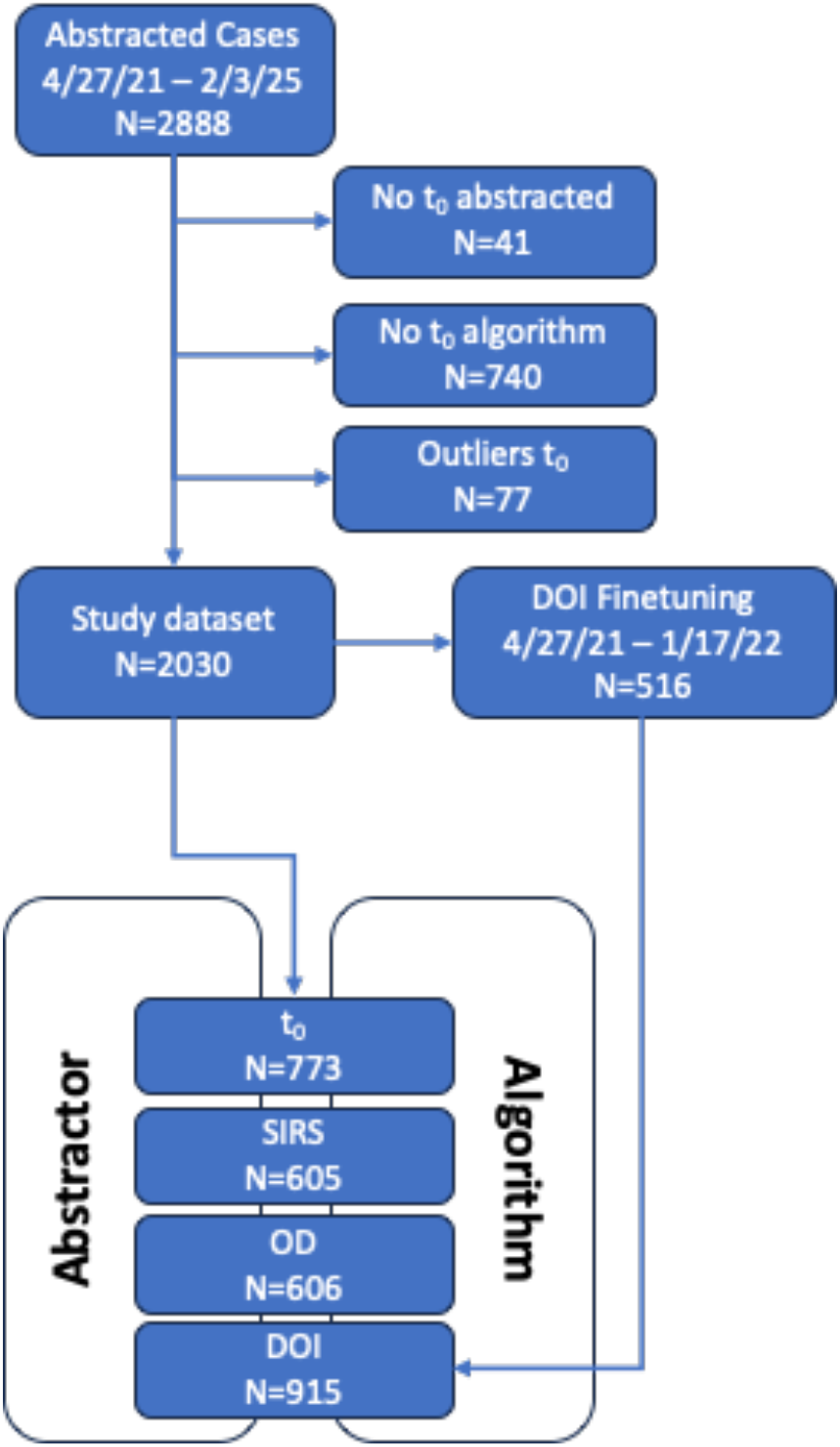
Consort diagram.

**Figure 2.**
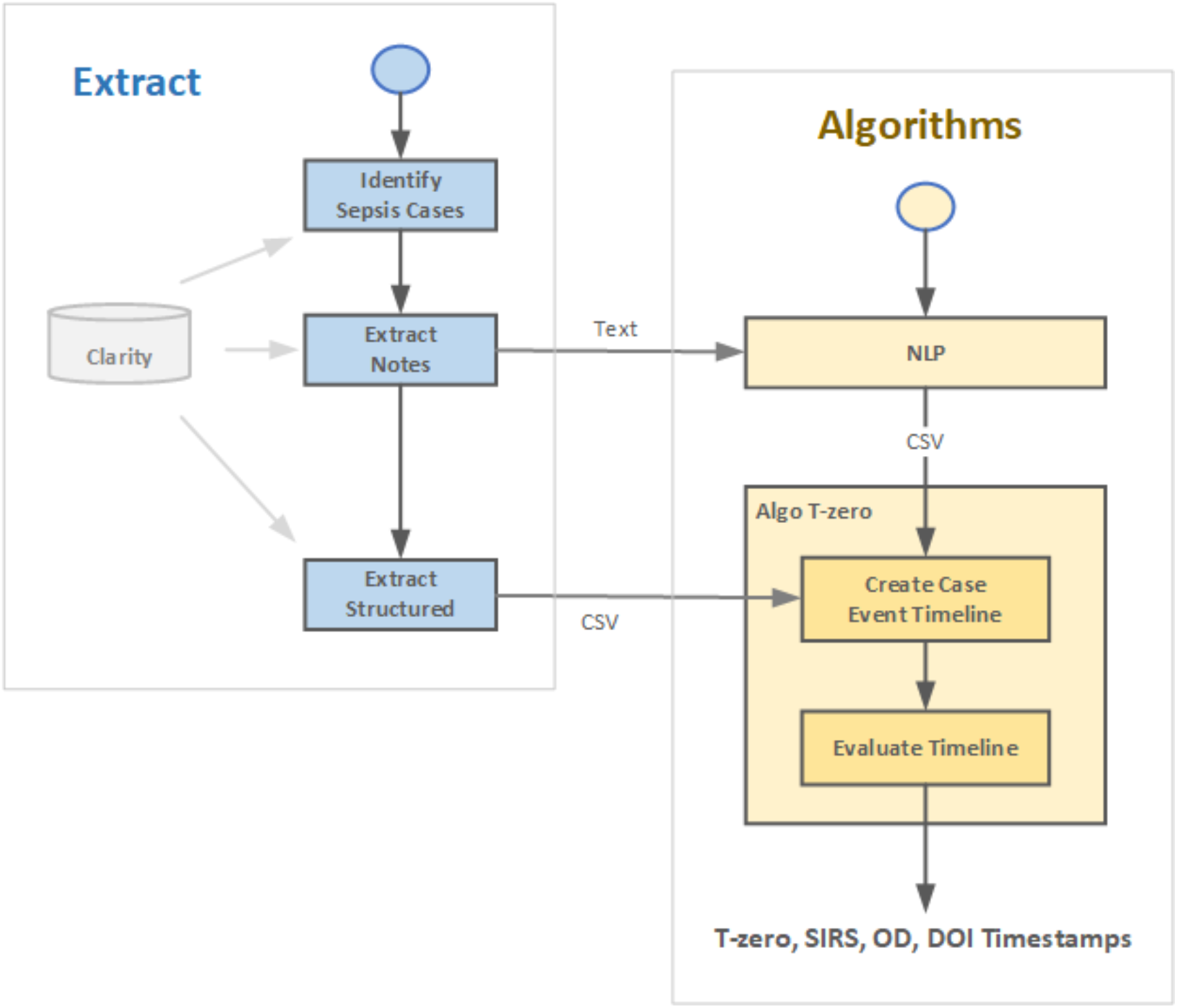
Workflow diagram of the sepsis algorithm.

**Figure 4.**
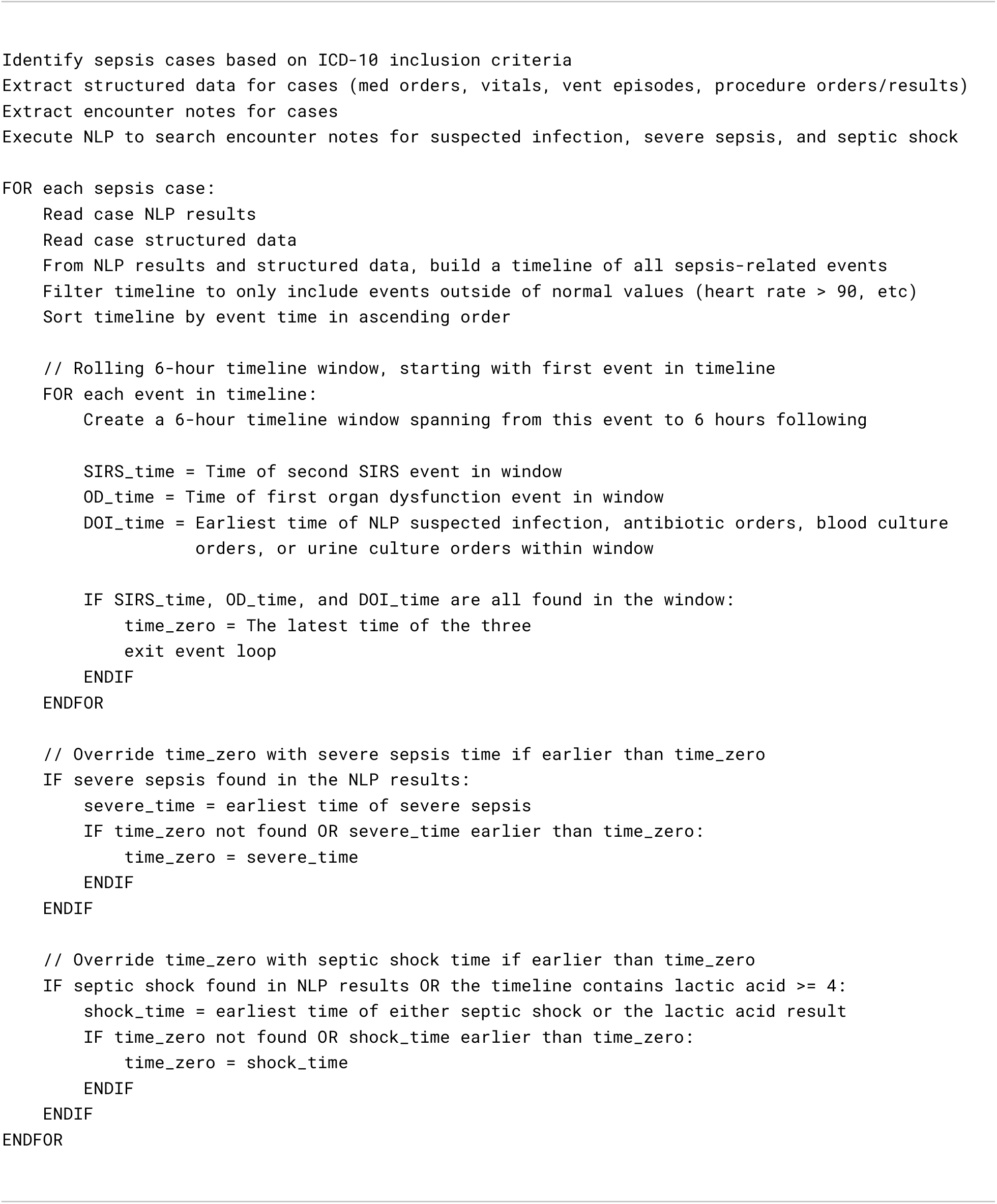
Sepsis t_0_ pseudocode.

**Figure 5.**
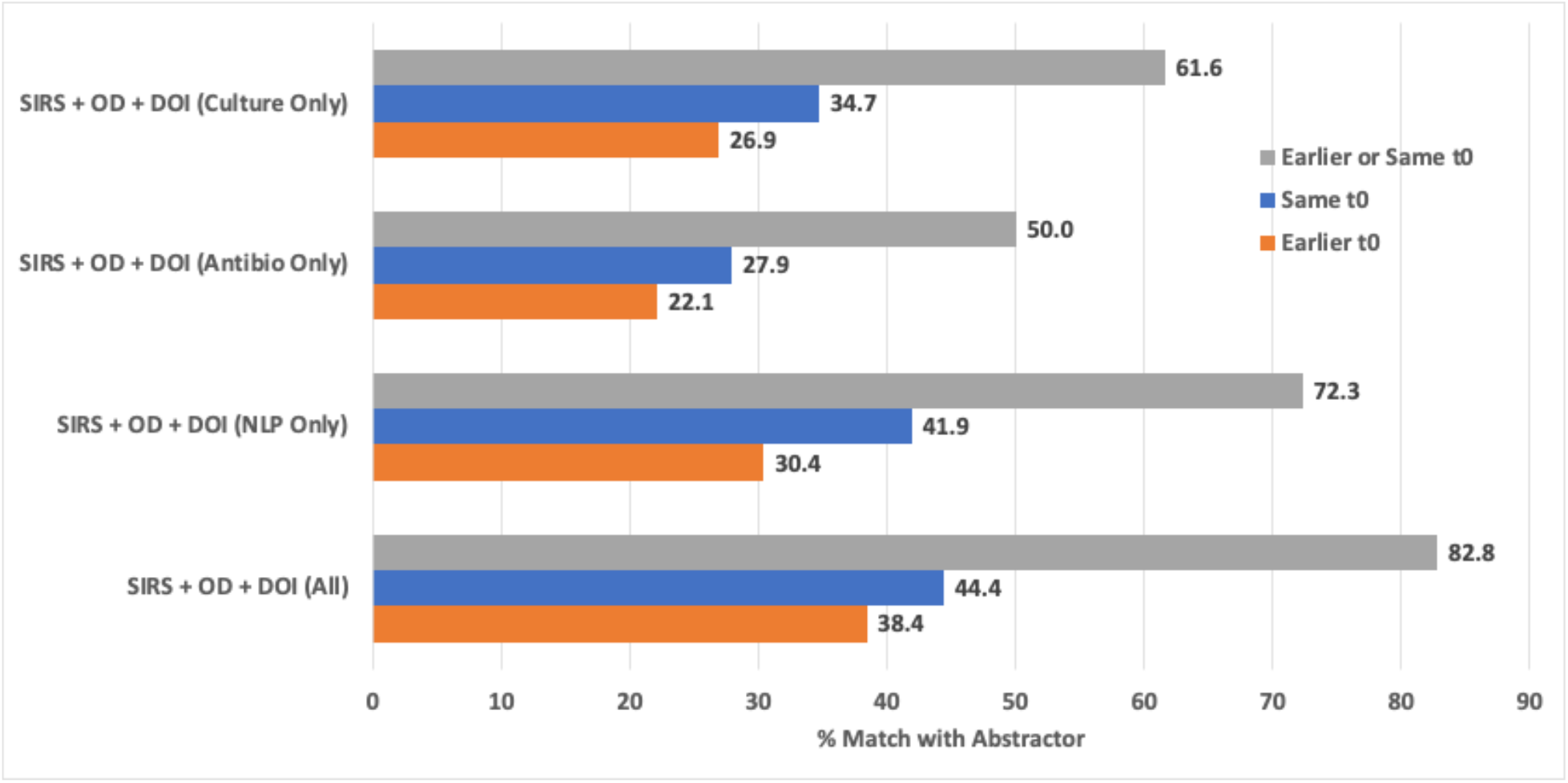
Precision of t_0_ algorithm using different DOI definitions.

**Figure 6.**
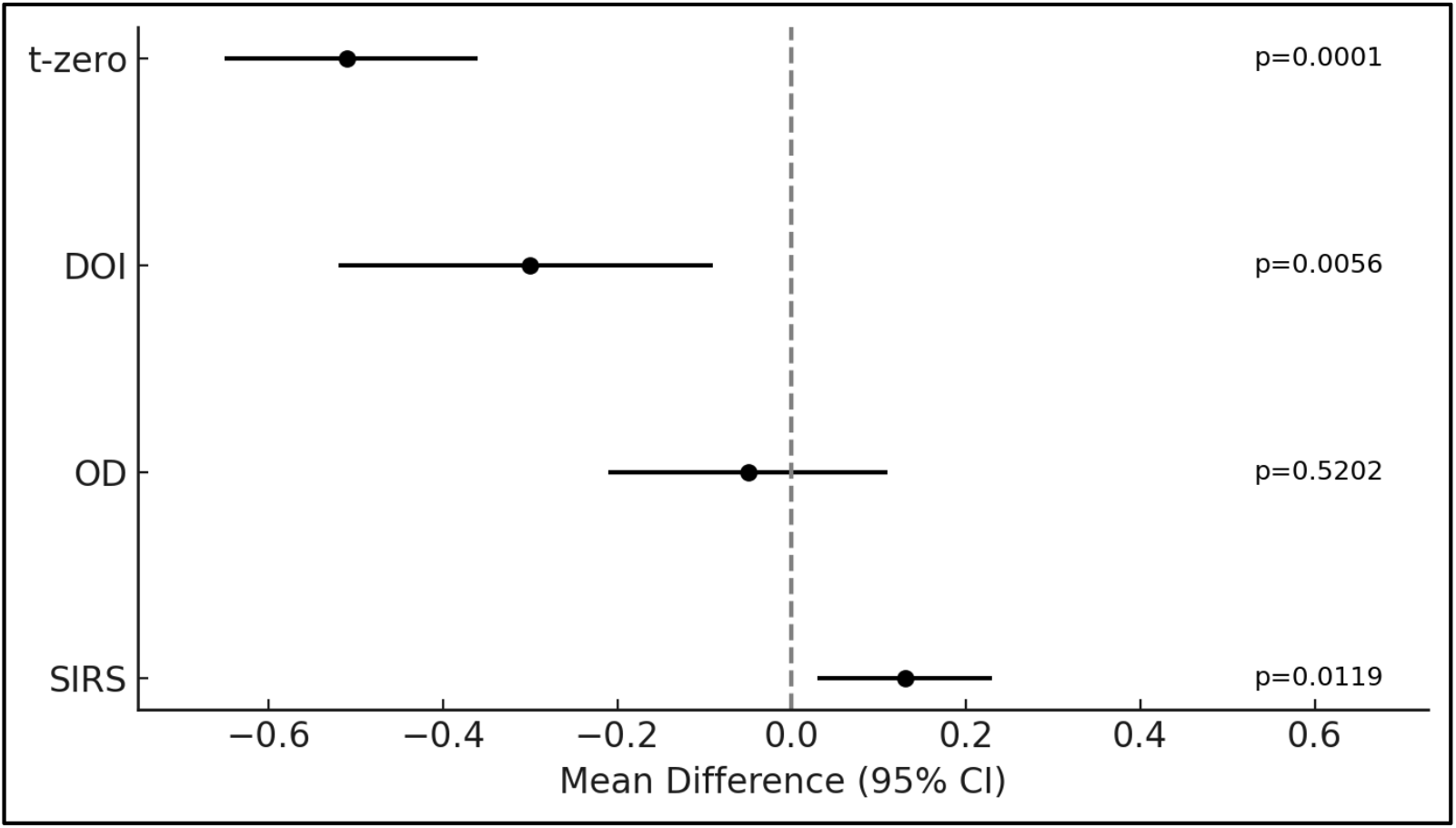
Differences between algorithm and abstractor defined sepsis t_0_ and its components.

Automated, note-aware identification of t_0_ also enables larger-scale, more granular studies of sepsis epidemiology and outcomes. By anchoring encounters at a clinically relevant t_0_, researchers can more reliably examine how patient factors, care processes, and system features influence trajectories and outcomes across diverse populations and settings. Automated extraction of t_0_ and related bundle elements can further support near real-time surveillance, feedback, and targeted training as emphasized in national guidance for hospital sepsis programs.^1^

The algorithm did not significantly differ from abstractors in detecting organ dysfunction, but it did identify SIRS criteria later. This delay is largely attributable to emergency medical services (EMS) documentation of pre-hospital vitals into scanned documents, which are not accessible to the algorithm in its current form. Performance could be improved by incorporating this information; however, reliable extraction from scanned documents remains a technical challenge despite advances in optical character recognition (OCR) and natural language processing (NLP).^12^ Moreover, EMS data integration into hospital EMRs remains inconsistent and unreliable.^13^

Other limitations of our study include the reliance on rule-based regular expressions to identify suspicion of infection within clinical notes. Such approaches are inherently brittle and may not generalize well across different note types or documentation styles. In addition, our work was conducted within a single health system, which may limit generalizability. However, the dataset encompassed eight hospitals in distinct geographic locations and diverse patient populations, partially mitigating this concern. Future research should evaluate the transferability of the NLP algorithm across other health systems and directly compare its performance with large language models, which are expected to offer greater flexibility in detecting suspicion of infection from physician notes.

## Conclusions

Our algorithm integrates structured data and clinical notes to identify sepsis t_0_ more precisely than proxies alone. Overall, it detected DOI significantly earlier than human abstractors. Scalable automation can reduce abstractor burden and human error, strengthen sepsis surveillance, and enable robust research to improve patient care.

## Data Availability

The source data for this study cannot be shared due to privacy concerns related to unstructured clinical notes. However, the Regular Expressions used for data extraction are available from the authors upon reasonable request.

## Acknowledgements

We are grateful for the support provided by the Corewell Health Research Institute and by Oakland University William Beaumont School of Medicine. We are also grateful for the valuable insights and feedback provided by the Corewell Health East Quality and Safety team during the development of the algorithm and for providing the abstracted cases used for benchmarking the algorithm.

## Author Contributions

Ramin Homayouni (Conceptualization, Formal Analysis, Project Administration, Writing - original draft), Joel Karsten (Writing - original draft), Riya Chahbra (Writing - original draft), Shane Morrell (Data curation, Methods, Formal Analysis, Writing - original Draft), Paul Bozyk (Conceptualization, Writing - review & editing).

## SUPPLEMENTARY MATERIALS

**Figure S1:**
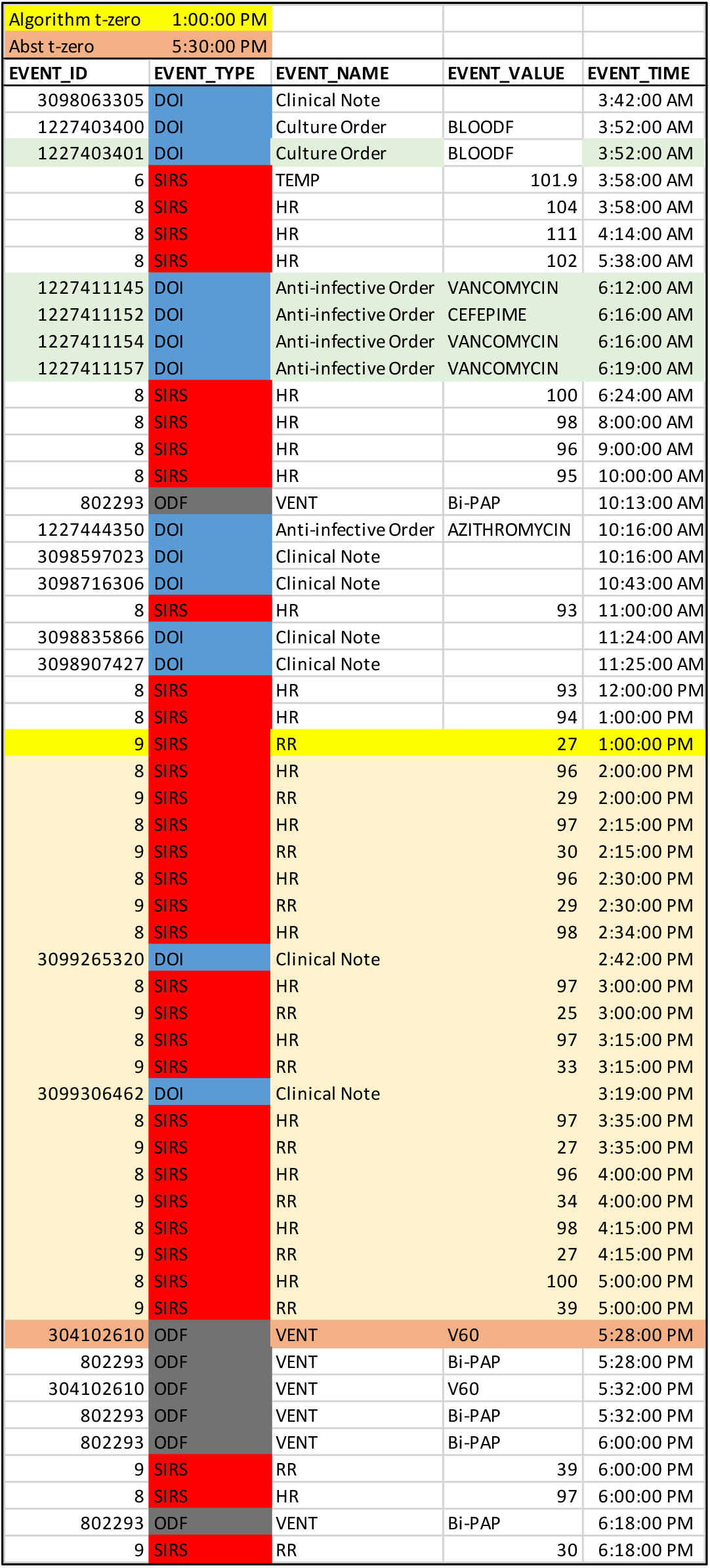
Sample Patient Timeline. The algorithm event log for DOI, SIRs, and OD after arrival to the hospital. The algorithm t_0_ was 13:00, whereas the abstractor t_0_ was 17:30.

